# Risk factors for severity on admission and the disease progression during hospitalization in a large cohort of COVID-19 patients in Japan

**DOI:** 10.1101/2021.04.02.21254809

**Authors:** Mari Terada, Hiroshi Ohtsu, Sho Saito, Kayoko Hayakawa, Shinya Tsuzuki, Yusuke Asai, Nobuaki Matsunaga, Satoshi Kutsuna, Wataru Sugiura, Norio Ohmagari

## Abstract

**Objectives:** To investigate the risk factors contributing to severity on admission. Additionally, risk factors on worst severity and fatality were studied. Moreover, factors were compared based on three points: early severity, worst severity, and fatality.

**Design:** A observational cohort study utilizing data entered in a Japan nationwide COVID-19 inpatient registry, COVIREGI-JP.

**Setting:** As of August 31, 2020, 7,546 cases from 780 facilities have been registered. Participating facilities cover a wide range of hospitals where COVID-19 patients are admitted in Japan.

**Participants:** Participants who had a positive test result on any applicable SARS-CoV-2 diagnostic tests, and were admitted to participating healthcare facilities. A total of 3,829 cases were identified from January 16 to May 31, 2020, of which 3,376 cases were included in this study.

**Primary and secondary outcoe measures:** Primary outcome was severe or non-severe on admission, determined by the requirement of mechanical ventilation or oxygen therapy, SpO2, or respiratory rate. Secondary outcome was the worst severity during hospitalization, judged by the requirement of oxygen and/or IMV/ECMO.

**Results:** Risk factors for severity on admission were older age, male, cardiovascular disease, chronic respiratory disease, diabetes, obesity, and hypertension. Cerebrovascular disease, liver disease, renal disease or dialysis, solid tumor, and hyperlipidemia did not influence severity on admission; however it influenced worst severity. Fatality rates for obesity, hypertension, and hyperlipidemia were relatively lower.

**Conclusions:** This study segregated the comorbidities driving severity and death. It is possible that risk factors for severity on admission, worst severity, and fatality are not consistent and may be propelled by different factors. Specifically, while hypertension, hyperlipidemia, and obesity had major effect on worst severity, their impact was mild on fatality in the Japanese population. Some studies contradict our results; therefore, detailed analyses, considering in-hospital treatments, are needed for validation.

**Trial registration:** UMIN000039873. https://upload.umin.ac.jp/cgi-open-bin/ctr_e/ctr_view.cgi?recptno=R000045453

**Strengths and limitations of this study:**

- In this article, we studied the disease progression of COVID-19, by comparing the risk factors on three points: early severity, worst severity, and fatality.
- Our results are useful from a public health perspective, as we provide risk factors for predicting the severity on admission and disease progression from patients’ background factors.
- This study pointed out the possibility that risk factors of the severity on admission, worst severity, and fatality are not consistent and may be propelled by different factors.
- Our data were collected from hundreds of healthcare facilities; thus data accuracy may be questionable.
- Also, treatment type, dosage, duration, and combination varied immensely across the facilities and we did not consider treatments prior to and during hospitalization in the analysis.

## Introduction

Coronavirus disease 2019 (COVID-19), caused by severe acute respiratory syndrome coronavirus-2 (SARS-CoV-2), has caused a major global public health crisis. As of October 3, 2020, >34 million people had been infected in over 230 countries^1, 2^. Japan experienced two pandemic waves after the first case reported on January 16, 2020. During the first wave, a state of emergency was declared on April 7, which ended on May 25, settling the first wave. Nearly thrice as many SARS-CoV-2 positive cases were detected in the second wave, which emerged from the end of June^3^. The fatality rate in the second wave has generally been lower in many countries, including Japan^4^.

When the number of patients explodes, hospital beds were in great shortage; hotels were utilized as isolation facilities in many countries^5–7^. Likewise, in Japan, mild patients were transferred to hotels from April 2020^8^. About two-thirds of cases did not require oxygen support throughout thier illness^9^. However, some cases initiated non-severe may instantly plunge into a serious state and require aggressive care^10^. Therefore, public health centers are in demand for indicators to identify those at a higher risk of aggravation in the early phase and determine the destination—hospital, hotel, or home. Depicting the clinical course—from onset to worst severity and the outcome—is imperative to appropriately allocate patients to healthcare resources. Analyses considering the severity on admission and the disease progression thereafter has not been conducted are of interest to physicians globally.

We obtained nationwide data from a COVID-19 inpatient registry, “COVID-19 REGISTRY JAPAN (COVIREGI-JP)”^11^, and conducted a study to identify the independent risk factors contributing towards severity on admission. We aimed to determine the risk factors on admission, namely demographics and comorbidities. Progression of severity was inspected in detail on different time points. Cases identified within the period of the first pandemic wave were studied.

## Methods

### Study design and patients

This is an observational cohort study that utilizes the data accumulated in the nationwide “COVID-19 REGISTRY JAPAN (COVIREGI-JP)”^11^. As of September 28, 2020, 10,048 cases from 802 facilities have been registered. Participating facitilies covers a wide range of hospitals where COVID-19 patients are admitted in Japan. Enrolled cases satisfied two eligibility criteria: a positive test result for COVID-19 and being admitted to a healthcare facility. Registration started on March 2, 2020, and is ongoing, at present.

### Data collection and case report form

Data were collected in a case report form (CRF) developed for COVIREGI-JP. This CRF includes modified information of the International Severe Acute Respiratory and Emerging Infection Consortium CRF on COVID-19^12^. Upon modification, we elaborated on data collection, especially on treatments, comorbidities, and symptoms. In addition, as of October 26, 2020, this CRF underwent revisions twice to update therapeutic options or definitions, as new evidence emerges. Study data were collected and managed using Research Electronic Data Capture (REDCap) electronic data capture tools^13, 14^, hosted at the datacenter in National Center for Global Health and Medicine. Data were either recorded on a CRF hard-copy or were entered directly into REDCap at each facility.

### Comorbidities

Comorbidities were collected based on Charlson comorbidity index^15, 16^ with modifications. Some comorbidities were combined as follows: Cardiovascular disease (CVD)—myocardial infarction, congestive heart failure, and peripheral vascular disease; Chronic respiratory disease (CRD)—chronic obstructive pulmonary disease (COPD) and other chronic lung diseases; Renal disease or dialysis—moderate to severe renal disorder (creatinine ≥ 3mg/dL, nephropathy, post-renal transplantation, or on dialysis), and maintenance hemodialysis or peritoneal dialysis before hospitalization; and Solid tumor—solid tumor with or without metastasis. Obesity was diagnosed based on physician’s judgement, and body mass index (BMI) was not considered in this study.

### Drug administration prior to and during hospitalization

Steroids, chemotherapy, and immunosuppressants administered prior to hospitalization were collected as pre-hospitalization treatments. Steroids included those equivalent to 20 mg/day prednisolone for ≥1 month and are not considered as immunosuppressants. Chemotherapy and immunosuppressants was applicable if administered 3 months prior to hospitalization. Treatment during hospitalization was studied on systemic steroids, favipiravir, ciclesonide, heparin, and tocilizumab, due to the frequent use in Japan. Heparin use included those given for both prophylactic and treatment purposes.

### Dataset

We defined the first wave period as January 16 to May 31, 2020^17^, and cases from the first wave was included in this analysis. Therefore, data extraction conditions were: (1) cases admitted to healthcare facilities between January 16 and May 31, 2020, and (2) all CRF items completed on dataset generation. The dataset was generated and fixed on September 2, 2020.

### Definitions of severity

#### 1) Severity on admission

Severity on admission was converted into bivariate variables: severe and non-severe. Cases met at least one of the following criteria were categorized as severe: (1) requiring invasive or non-invasive mechanical ventilation, (2) requiring supplemental oxygen, (3) SpO_2_ ≤94% in room air, or (4) tachypnea with respiratory rate (RR) ≤24 breaths per minute^18^. Those who did not meet the aforementioned were classified as non-severe.

#### 2) Worst severity

The worst severity was grouped into three categories: no-oxygen, oxygen, and IMV/ECMO. The worst state during hospitalization was adopted on categorization, and each was defined as follows:

No-oxygen—No requirement of supplemental oxygen throughout hospitalization. Oxygen—Required supplemental oxygen (including high-flow oxygen devices) or non-invasive mechanical ventilation during hospitalization.

IMV/ECMO—Required invasive mechanical ventilation or extracorporeal membrane oxygenation during hospitalization.

### Statistical analysis

Continuous variables are presented in median and interquartile range (IQR) and categorical variables in number of cases and percentages. We classified the disease progression into three stages: severity on admission, worst severity, and clinical outcomes. We used Mann-Whitney U tests (for two groups) or Kruskal-Wallis tests (for three groups) for continuous variables and chi-squared tests for categorical variables.

We conducted a multivariable logistic regression analysis to identify the factors associated with the patients’ severity on admission. We included age, sex, comorbidities (CVD, cerebrovascular disease, CRD, asthma, liver disease, diabetes, obesity diagnosed by physicians, renal disease or dialysis, solid tumors, leukemia, lymphoma, hypertension, and hyperlipidemia), use of systemic steroids in the past month, chemotherapy in the past three months, and use of immunosuppressants other than steroids as independent variables. Multicollinearity was evaluated using the variance inflation factor (VIF). Variables of VIF > 3 were excluded from the model; however, no variables demonstrated obvious multicollinearity.

R version3.6.3 (R core team, 2020)^19^ was used for all the analyses performed in this study.

### Ethics

The National Center for Global Health and Medicine ethics board approved this study (referral number NCGM-G-003494-08), and waived the need for informed consent from individual patients owing to the non-invasive, non-interventional nature of this observational study according to the local Ethical Guidelines^20^. Information regarding opting out of our study is available on the website^11^. Although it is not mandatory, the study is also being registered on trial registration website (Unique ID: UMIN000039873, https://upload.umin.ac.jp/cgi-open-bin/ctr_e/ctr_view.cgi?recptno=R000045453).

## Results

Within the study period, 3,829 cases were identified and 3,376 cases from 299 facilities were included in this study. Of them, 2,199 cases (65.1%) were non-severe, and 1,181 cases (34.9%) were severe at the time of admission. After categorizing the two groups further into no-oxygen, oxygen, and IMV/ECMO by worst severity, compositions were 1,758 (81.5 %), 357 (16.5%), and 43 (2.0%) for the non-severe group and 190 (16.1%), 677 (57.5%), and 311 (26.4%) for the severe group, respectively. While categorizing the cases, 44 (1.3%) were unavailable due to missing values.

Demographics and clinical characteristics of the study population are shown in Table 1. Days between onset and admission were similar in both groups (non-severe 6.0 vs severe 7.0 days). Over ten times as many severe cases on admission underwent IMV/ECMO than non-severe cases (2.0% vs 26.4%). Severe cases were older (50.0 vs 67.0), had higher BMI (22.9 vs 24.1), greater male dominance (56.3% vs 70.5%), and a higher prevalence of comorbidities excluding leukemia, compared to the non-severe group. The most prevalent symptoms in both groups were fever (non-severe 49.5%, severe 73.7%), cough (non-severe 53.8%, severe 64.9%), and fatigue (non-severe 40.3%, 59.9%), but was greater in the severe group. Conversely, prevalence of dysgeusia (25.9% vs 13.2%), dysosmia (22.6% vs 11.5%), headache (18.1% vs 14.7%), and runny nose (11.9% vs 8.9%) was higher in the non-severe group.

**Table 1.** Characteristics of patients included in the present study.

|  | Non-severe (n=2196) | Severe (n=1180) |
| --- | --- | --- |
| **Fatal cases** | 39 (2) | 204 (17) |
| **Worst severity during hospitalization** |  |  |
| **No oxygen** | 1796 (82) | 192 (16) |
| **Oxygen** | 357 (16) | 678 (58) |
| **IMV/ECMO<sup>a</sup>** | 43 (2) | 310 (26) |
| **Days between onset and admission**<br>**(median [IQR])** | 6 [4, 10] | 7 [4, 10] |
| **Age (median [IQR])** | 50 [35, 64] | 67 [53, 78] |
| **Male** | 1232 (56) | 830 (71) |
| **BMI (median [IQR])** | 22.9 [20.3, 25.7] | 24.1 [21.5, 27.1] |
| **Comorbidities** |  |  |
| **Cardiovascular disease** | 62 (3) | 121 (10) |
| **Respiratory disease** | 36 (2) | 104 (9) |
| **Liver disease** | 49 (2) | 39 (3) |
| **Cerebrovascular disease** | 72 (3) | 115 (10) |
| **Asthma** | 102 (5) | 64 (5) |
| **Diabetes** | 262 (12) | 300 (25) |
| **Obesity** | 95 (4) | 83 (7) |
| **Severe renal disease or dialysis** | 22 (1) | 25 (2) |
| **Solid tumor** | 66 (3) | 79 (7) |
| **Leukemia** | 10 (1) | 3 (0) |
| **Lymphoma** | 16 (1) | 9 (1) |
| **Hypertension** | 292 (13) | 331 (28) |
| **Hyperlipidemia** | 176 (8) | 157 (13) |
| **Treatments prior to COVID-19** |  |  |
| **Use of steroid in one month** | 6 (0) | 10 (1) |
| **Chemotherapy in three months** | 32 (2) | 24 (2) |
| **Immunosuppressants<sup>b</sup> use in three months** | 26 (1) | 18 (2) |
| **Symptoms on admission** |  |  |
| **Fever ($\geq 37.5^{\circ}\text{C}$)** | 1078 (49) | 862 (74) |
| **Cough** | 1167 (54) | 716 (65) |
| **Sore throat** | 340 (17) | 142 (16) |
| **Runny nose** | 239 (12) | 86 (9) |
| **Chest pain** | 95 (5) | 44 (5) |
| **Myalgia** | 172 (9) | 79 (9) |
| **Headache** | 361 (18) | 136 (15) |
| **Confusion** | 21 (1) | 68 (6) |
| **Fatigue** | 834 (40) | 595 (60) |
| **Abdominal pain** | 60 (3) | 24 (3) |
| **Vomit** | 88 (4) | 59 (6) |
| **Diarrhea** | 251 (12) | 164 (16) |
| **Dysgeusia** | 494 (26) | 113 (13) |
| **Dysosmia** | 422 (23) | 96 (12) |

<sup>a</sup>invasive mechanical ventilation/extracorporeal membrane oxygenation
<sup>b</sup>immunosuppressants other than steroids

Results of the multivariate logistic regression to determine the risk of severity on admission are shown in Table 2. Older age (OR 1.038 [1.032—1.044]) and male (OR 2.06 [1.69—2.51]) were considered a risk among the demographics and comorbidities included CVD (OR 1.61 [1.07—2.43]), respiratory disease (OR 2.59 [1.63—4.13]), diabetes (OR 1.39 [1.09—1.76]), obesity (OR 1.62 [1.12—2.35]), and hypertension (OR 1.31 [1.03—1.67]). Days between onset to admission were non-significant (*p* = 0.376); the timing of admission did not affect the severity on admission.

**Table 2.** Factors associated with being “severe” at the time of admission.

|  | **Odds ratio** | **95% CI<sup>a</sup>** | **P value** |
| --- | --- | --- | --- |
| **Days between onset and admission** | 1.0 | 0.99-1.01 | 0.960 |
| **Age** | 1.04 | 1.03-1.04 | < 0.001 |
| **Male** | 2.09 | 1.76-2.48 | < 0.001 |
| **Comorbidities** |  |  |  |
| **Cardiovascular disease** | 1.48 | 1.04-2.10 | 0.028 |
| **Cerebrovascular disease** | 1.33 | 0.95-1.85 | 0.097 |
| **Chronic respiratory disease** | 2.51 | 1.67-3.78 | < 0.001 |
| **Asthma** | 1.24 | 0.87-1.77 | 0.240 |
| **Liver disease** | 0.97 | 0.61-1.54 | 0.892 |
| **Diabetes** | 1.34 | 1.09-1.64 | 0.006 |
| **Obesity diagnosed by physicians** | 1.75 | 1.26-2.45 | 0.001 |
| **Severe renal disease or dialysis** | 1.0 | 0.54-1.88 | 0.991 |
| **Solid tumor** | 1.20 | 0.82-1.77 | 0.351 |
| **Leukemia** | 0.34 | 0.08-1.39 | 0.132 |
| **Lymphoma** | 0.42 | 0.16-1.11 | 0.081 |
| **Hypertension** | 1.33 | 1.08-1.64 | 0.008 |
| **Hyperlipidemia** | 0.91 | 0.70-1.19 | 0.490 |
| **Treatments prior to COVID-19** |  |  |  |
| **Use of steroid in one month** | 1.65 | 0.52-5.22 | 0.394 |
| **Chemotherapy in three months** | 1.47 | 0.72-3.0 | 0.286 |
| **Immunosuppressants<sup>b</sup> use in three months** | 1.35 | 0.69-2.64 | 0.384 |

<sup>a</sup>confidence interval
<sup>b</sup>immunosuppressants other than steroids

Table 3 depicts the study population from a different angle and is categorized by the worst severity (n=3,336) and fatality (n=3,376). Oxygen and IMV/ECMO cases were predominantly severe at admission (65.5% and 87.9%, respectively), whereas most no-oxygen cases come from non-severe group (90.2%). Prevalence of comorbidities was lowest in no-oxygen cases; however, prominent difference was not observed for asthma. Similarly, fatal cases were more severe at admission (84.0% *vs* 31.1%) and had higher prevalence of oxygen and IMV/ECMO cases (oxygen: 56.4% *vs* 29.0%, IMV/ECMO: 41.9% *vs* 8.2%, respectively). Days between onset and admission was longer in non-fatal cases (5-days *vs* 7-days).

**Table 3.** Characteristics of patients stratified by non-fatal/fatal cases and severity during hospitalization.

|  | **Non-fatal**<br>**(n=3129)** | **Fatal**<br>**(n=243)** | **No-oxygen**<br>**(n=1988)** | **Oxygen**<br>**(n=1035)** | **IMV/ECMO<sup>a</sup>**<br>**(n=353)** |
| --- | --- | --- | --- | --- | --- |
| **Fatal cases** |  |  | 6 (0) | 137 (13) | 100 (28) |
| **Severity on admission** |  |  |  |  |  |
| **Non-severe** | 2155 (69) | 39 (16) | 1796 (90) | 357 (35) | 43 (12) |
| **Severe** | 974 (31) | 204 (84) | 192 (10) | 678 (66) | 310 (88) |
| **Worst severity during hospitalization** |  |  |  |  |  |
| **No-oxygen** | 1980 (63) | 6 (3) |  |  |  |
| **Oxygen** | 897 (29) | 137 (56) |  |  |  |
| **IMV/ECMO** | 252 (8) | 100 (41) |  |  |  |
| **Days between onset and admission (median [IQR])** | 7 [4, 10] | 5 [2, 8] | 7 [4, 10] | 6 [3, 9] | 7 [5, 10] |
| **Age (median [IQR])** | 54 [40, 68] | 80 [71, 86] | 48 [33, 61] | 68 [53, 80] | 65 [56, 74] |
| **Male** | 1899 (61) | 161 (66) | 1083 (55) | 694 (67) | 285 (81) |
| **BMI (median [IQR])** | 23.3 [20.8,<br>26.3] | 22.7 [19.4,<br>25.7] | 22.6 [20.2,<br>25.5] | 24.0<br>[21.5,<br>27.0] | 24.8 [22.6,<br>27.8] |
| **Cardiovascular disease** | 129 (4) | 54 (22) | 48 (2) | 106 (10) | 29 (8) |
| **Respiratory disease** | 103 (3) | 35 (14) | 29 (2) | 78 (8) | 33 (9) |
| **Liver disease** | 75 (2) | 13 (5) | 36 (2) | 32 (3) | 20 (6) |
| **Cerebrovascular disease** | 135 (4) | 51 (21) | 57 (3) | 105 (10) | 25 (7) |
| **Asthma** | 157 (5) | 9 (4) | 92 (5) | 52 (5) | 22 (6) |
| **Diabetes** | 475 (15) | 86 (35) | 197 (10) | 244 (24) | 121 (34) |
| **Obesity** | 169 (5) | 9 (4) | 70 (4) | 75 (7) | 33 (9) |
| **Severe renal disease or**<br>**dialysis** | 34 (1) | 13 (5) | 14 (1) | 21 (2) | 12 (3) |
| **Solid tumor** | 114 (4) | 31 (13) | 60 (3) | 63 (6) | 22 (6) |
| **Leukemia** | 9 (0) | 4 (2) | 6 (0) | 7 (1) | 0 (0) |
| **Lymphoma** | 13 (0) | 12 (5) | 6 (0) | 16 (2) | 3 (1) |
| **Hypertension** | 551 (18) | 70 (29) | 234 (12) | 274 (27) | 115 (33) |
| **Hyperlipidemia** | 305 (10) | 26 (11) | 148 (7) | 124 (12) | 61 (17) |
| **Use of steroid in one** | 10 (0) | 5 (2) | 4 (0) | 8 (1) | 4 (1) |

| month |  |  |  |  |  |
| --- | --- | --- | --- | --- | --- |
| **Chemotherapy in three months** | 38 (1) | 18 (7) | 21 (1) | 30 (3) | 5 (1) |
| **Immunosuppressants<sup>b</sup> use in three months** | 37 (1) | 7 (3) | 18 (1) | 18 (2) | 8 (2) |
| **Fever ($\geq 37.5^{\circ}\text{C}$)** | 1737 (56) | 199 (82) | 897 (45) | 758 (74) | 285 (82) |
| **Cough** | 1770 (58) | 112 (52) | 1034 (53) | 643 (64) | 206 (69) |
| **Sore throat** | 459 (17) | 23 (16) | 316 (18) | 125 (15) | 41 (16) |
| **Runny nose** | 311 (11) | 13 (7) | 224 (12) | 79 (9) | 22 (8) |
| **Chest pain** | 136 (5) | 3 (2) | 88 (5) | 46 (6) | 5 (2) |
| **Myalgia** | 242 (9) | 9 (7) | 148 (8) | 84 (10) | 19 (7) |
| **Headache** | 486 (18) | 11 (8) | 333 (18) | 138 (17) | 26 (10) |
| **Confusion** | 60 (2) | 29 (14) | 19 (1) | 54 (6) | 16 (5) |
| **Fatigue** | 1323 (46) | 104 (62) | 709 (38) | 560 (62) | 160 (58) |
| **Abdominal pain** | 79 (3) | 5 (3) | 53 (3) | 25 (3) | 6 (2) |
| **Vomit** | 139 (5) | 8 (5) | 80 (4) | 51 (6) | 16 (6) |
| **Diarrhea** | 397 (14) | 18 (9) | 236 (13) | 143 (15) | 36 (13) |
| **Dysgeusia** | 128 (17) | 25 (10) | 454 (26) | 128 (17) | 25 (10) |
| **Dysosmia** | 103 (14) | 13 (6) | 402 (23) | 103 (14) | 13 (6) |

<sup>a</sup>invasive mechanical ventilation/extracorporeal membrane oxygenation
<sup>b</sup>immunosuppressants other than steroids

More non-severe cases with any comorbidity underwent treatment with oxygen or IMV/ECMO compared to non-severe cases with no comorbidities. In figure 1, only 11.9% underwent oxygen therapy or IMV/ECMO in non-severe cases without any comorbidities. However, among the non-severe cases with comorbidity, the rates of oxygen or IMV/ECMO were higher in most comorbidities, including CVD (34.7%), CRD (38.9%), liver disease (36.7%), diabetes (35.1%), obesity (35.8%), cerebrovascular disease (34.7%), renal disease (40.9%), solid tumor (27.3%), hypertension (31.2%), and hyperlipidemia (25.0%). Asthma alone followed a different trend; the chances of oxygen and IMV/ECMO requirement was lower.

**Figure 1.**
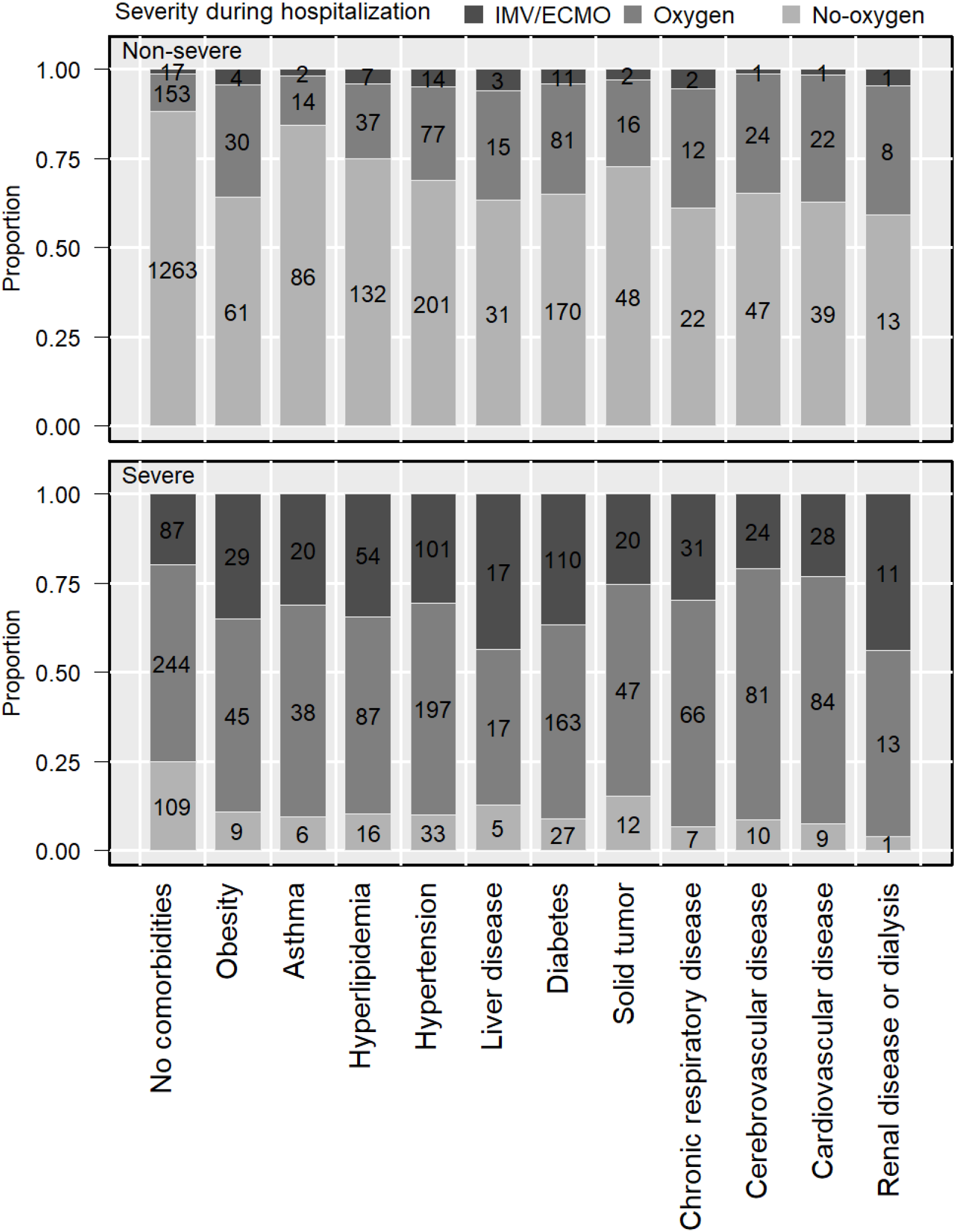
Distribution of the worst severity arranged by severe/non-severe at admission and presence/absence of comorbidities. Top bars represent non-severe cases at admission and bottom bars represent severe cases at admission. Each group of cases was divided based on the presence of comorbidities. Bars represent different categories of worst severity: light gray – no-oxygen, darker gray – oxygen, and darkest gray – IMV/ECMO.

Among the cases without comorbidity, 75.2% of cases that were severe on admission required oxygen or IMV/ECMO; however, the fatality rate was low, and only 8.0% resulted in death (Figure 2). Fatality rates were approximately 3—5 times higher when the following comorbidities were present: renal disease or dialysis (44%), CVD (40.5%), cerebrovascular disease (39.5%), CRD (30.4%), solid tumor (30.4%), diabetes (25.8%), and liver disease (25.6%). Even among non-severe cases, relatively high fatality rate was observed in cases with solid tumor, CRD, cerebrovascular disease, CVD, and renal disease or dialysis, with fatality rates ranging from 8.1% to 11.1%. Collectively, obesity, hypertension, and hyperlipidemia influenced the worst severity; however, their influence on fatality was relatively lower than that mentioned earlier.

**Figure 2.**
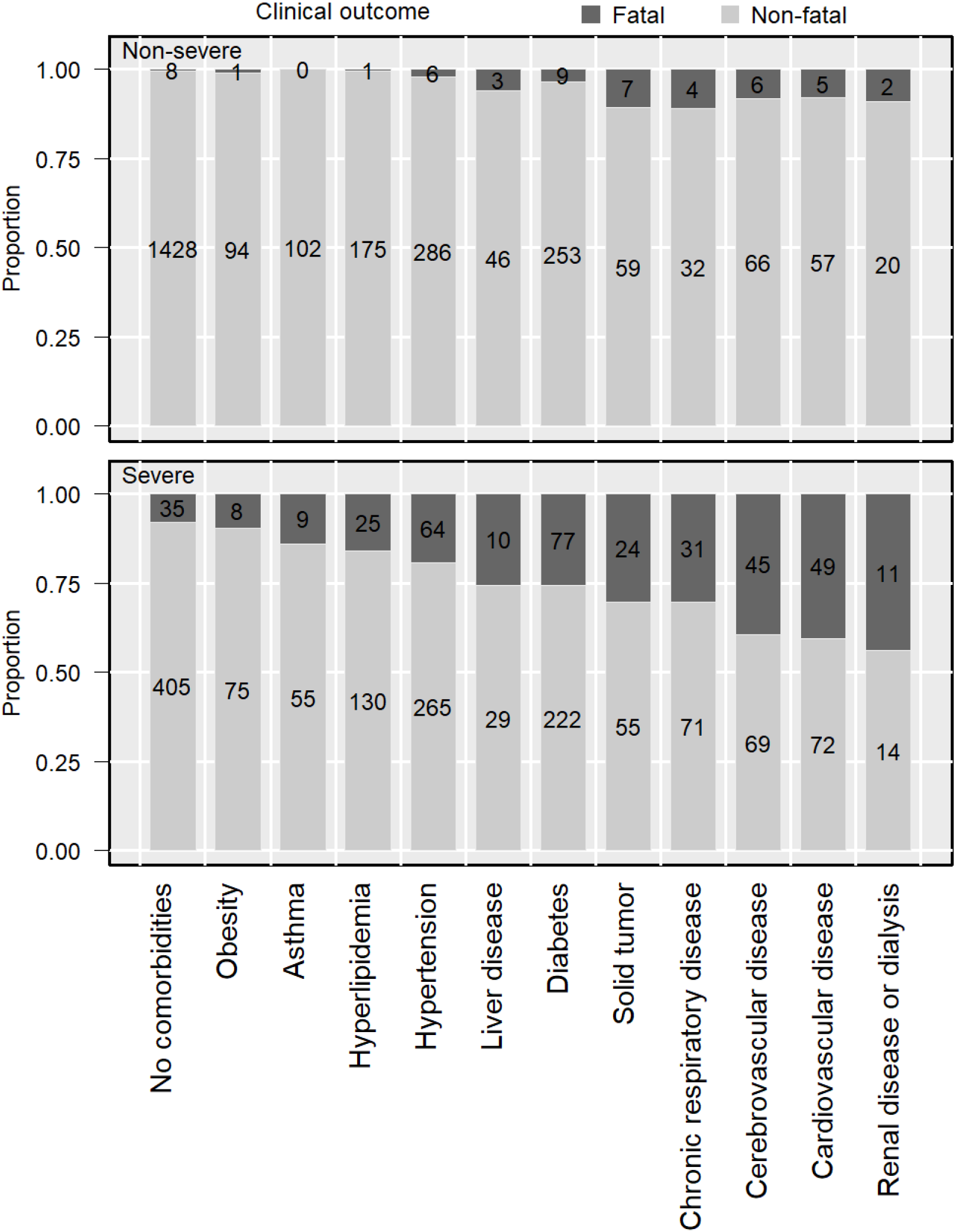
Distribution of the fatality arranged by severe/non-severe at admission and presence/non-presence of comorbidities. Top bars represent non-severe cases at admission and bottom bars represent severe cases at admission. Each group of cases was divided based on the presence of comorbidities. Dark gray represents fatal cases while light gray represents non-fatal cases.

Older age was relevant to both worst severity and fatality, as shown in supplemental figures 1 and 2. The combined proportion of oxygen and IMV/ECMO increased gradually by age from 5.3% in 20s to 69.3% in ≥80s. Conversely, the fatality rate leaped between 60s (2.2%) and 70s (8.6%). Likewise, supplemental figure 3 shows the combined proportion of oxygen and IMV/ECMO and fatality rates as higher in older individuals, irrespective of underlying comorbidities.

Predominant comorbid cases required more drug administration than those without comorbidities (Supplementary Table 1). Systemic steroids were most frequently used in cases with CRD (27.9%). Heparin was used most often in renal disease (12.8%), hypertension (11.2%), diabetes (10.9%), and CVD (10.4%).

## Discussion

We took disease progression into consideration and evaluated the study population based on severity on admission, worst severity, and the outcome. To our knowledge, studies have predominantly reported worst severity, whereas disease progression has not been considered. Our findings, therefore, are novel, augmenting the evidence needed to depict the clinical course and trajectory from onset to worsening condition. Specifically, this study segregated the comorbidities influencing severity and death. Based on our findings, it may be possible that the early severity, worst severity, and death are propelled by different factors, whilst confirmation is necessary by multivariate analysis.

The majority of comorbidities we studied did not influence severity on admission. On admission, severity was driven by age, sex, CVD, CRD, diabetes, obesity, and hypertension. The trend was similar for the worst severity, as cases with these factors had higher rate of oxygen or IMV/ECMO. However, all comorbidities appeared to influence the worst severity.

Within the comorbidities, the prognosis of cases with obesity, hypertension, or hyperlipidemia was relatively favorable. In contrast to our results, hypertension and obesity are reportedly related to an increased risk of severity and mortality^21–24^. However, a large cohort reported a trend similar to our results^25^. Another study reported that obesity is be confounded by age and sex^26, 27^. Obesity was judged by a physician in our study, and the results may change after incorporating BMI. Several other evidences suggest that the association of these comorbidities with poorer outcome of COVID-19 needs further investigation. BMI, on average, is lower in the Asia-Pacific region than in other global regions^28^; therefore, the degree of obesity may have been milder in our study population. Extreme obesity may worsen the prognosis, and confounders should be addressed in consecutive analyses. On the other hand, the presence of hypertension and the use of angiotensin-converting enzyme (ACE) inhibitors and angiotensin II receptor blockers act contrarily^29–31^, while ACE2 mediates the entry of SARS-CoV-2 into host cells^32, 33^, making the COVID-19 pathophysiology in hypertensive patients intricate. Our study suggested that hypertension, hyperlipidemia, and obesity could be less detrimental on fatality.

In accordance with previous studies, CVD, CRD, liver disease, diabetes, cerebrovascular disease, renal disease or dialysis, and solid tumor were associated with to fatality and worst severity. Two meta-analyses have reported common risk factors for worst severity during hospitalization, which include, diabetes, COPD, malignancy, CVD, and cerebrovascular disease^34, 35^. Other studies have also reported chronic liver disease and renal disease as risk factors^36–38^. Studies have elucidated that acute respiratory distress syndrome and coagulation dysfunction are related to the renin-angiotensin-aldosterone system and blood coagulation pathways, which are altered by SARS-CoV-2 host cell invasion via ACE2^39–41^. Clinical and non-clinical studies revealed an association between these comorbidities; while SARS-CoV-2 infection decreases ACE2 expression, ACE2 deficiency is reported to cause cardiac overload and kidney inflammation^41–44^. Elevated blood glucose is also associated with mortality^45^. Although risk factors vary among studies, the comorbidities we identified are highly likely associated with fatality, backed up by clinical and non-clinical results.

Different trends were seen in the rates of IMV/ECMO and death for each comorbidity. Although rates of IMV/ECMO were comparable in all comorbid cases, those with obesity, asthma, hyperlipidemia, and hypertension showed a lower fatality rate, suggesting that the fatality rates within the IMV/ECMO cases with these comorbidities were lower than expected. Contrarily, fatality rates in cases with CVD, cerebrovascular disease, renal dysfunction, tumor, and CRD were comparable or higher than IMV/ECMO rates. The number of death actually exceeded the number of IMV/ECMO cases in patients with tumor, cerebrovascular disease, or CVD. These comorbidities likely have caused a higher risk of death and some even died without intubation. Healthcare nearly overwhelmed in the first wave in Japan but ICU capacity was maintained^46^, and thus intubation may have been unperformed due to a medical judgment. A detailed examination of these issues is necessary in the future.

Our results did not show prominent difference in fatality between males and females. Oftentimes, males are considered to develop severe conditions and increased fatality^37, 38, 47–49^. However, according to Global Health 5050, sexual disparity in incidence of COVID-19 is low^50^. Additionally, ACE2 expression is affected by sexual hormones, whereby higher expression is observed in men, possibly explaining the sexual disparity^51–53^. Moreover, the immunological response to produce antibodies is more favorable in females^54^. These studies support the rationale that males are more susceptible to severe COVID-19, which contravene our results. The lower-than-expected fatality rate in our male population may be attributed to comorbidity prevalence, treatments, age, and/or degree of obesity.

Fatality rates were comparable between asthmatic and cases without comorbidities in our results. Theoretically, COVID-19 can be a risk for asthmatic patients. A viral respiratory infection is presented as relatively worse and causes asthma exacerbation^55, 56^. Asthmatic patients reportedly require a longer duration of mechanical ventilation when intubated^57^; however, no study, including ours, has found strong evidence on severity or mortality^58–61^. Inhaled corticosteroids (ICS) are known to downregulate ACE2^62, 63^ and are being investigated for treating COVID-19^64^. ICS may have impeded aggravation in asthmatic patients with COVID-19^65^. Overall, further studies are needed to elucidate the true risk of asthma on COVID-19.

Our results could be useful to roughly identify those at a risk of aggravation or death. Days from onset to admission was not a risk factor; early hospitalization will not influence the disease progression or outcome, and severity on admission was mostly driven by age and the presence of a few comorbidities. Several studies have created a scoring system to predict the risk of severity or mortality^66–68^. However, these utilize laboratory data collected on admission and are seldom practical for estimating the severity of illness prior to medical visits or when test results are not promptly available. While these are useful to predict prognosis more precisely, our results are useful from a public health perspective, as they provide risk factors for predicting the severity on admission and disease progression from patients’ background factors.

Our study has several limitations. In some of our analyses, confounders were not eliminated. We did not consider treatments prior to and during hospitalization. As our data were collected from hundreds of healthcare facilities, treatment type, dosage, duration, and combination varied immensely. We plan to deliberate the analytical methodology further to evaluate the outcomes which are prone to be affected by in-hospital treatments. Data were collected from numerous facilities; therefore, accuracy may be questionable. Additionally, hotels were utilized as isolation facilities from April 2020, and participant selection might have altered thereafter. COVIREGI-JP is continuously open for new entry; the number of registrations is increasing, and subsequent results may vary from ours.

## Conclusion

On admission, factors that influence severity were age, sex, and comorbidities, including CVD, CRD, diabetes, obesity, and hypertension. Risk factors for severity on admission, worst severity, and fatality were not consistent, and it is likely that they are each propelled by different factors. Our results are practically useful for predicting the progression and preparing for the worst, based on patients’ backgrounds. Moreover, based on our predictions, healthcare resources can be allocated to patients in the most suitable way.

## Data Availability

The data are not publicly available. The data set used was approved for the use in this study only.

## Acknowledgments

We thank all the participating facilities for their care towards COVID-19 patients and cooperation during data entry. We are especially grateful for the 299 facilities that contributed to the dataset used in this study.

## Contributorship statement

MT conceived and HO, SS, KH, ST and MT designed the study. ST, YA, SS, KH, and MT analyzed and interpreted the data. MT and ST drafted the first version of the manuscript. All the authors contributed to, read, and approved the final manuscript. The corresponding author attests that all listed authors meet authorship criteria and that no others meeting the criteria have been omitted. SS is the guarantor.

## Transparency Statement

The corresponding author affirms that this manuscript is an honest, accurate, and transparent account of the study being reported; that no important aspects of the study have been omitted; and that any discrepancies from the study as planned (and, if relevant, registered) have been explained.

## Data sharing

Data on an individual level is shared with limitation to participating healthcare facilities through applications to COVIREGI-JP^11^.

## Patient and Public Involvement

No patient was involved in the setting of research question, outcome measures, or study design, nor were they involved in the recruitment to and conduct of the study.

## Dissemination to participants and related patients and public communities

The study results will be shared with all the healthcare facilities which participated and registered data in COVIREGI-JP. It will also be shared with the public on the website^11^.

---

**Supplementary Table.** Proportion of therapeutics used for each comorbidity.

|  | **Number of patients** | **Systemic steroid** | **Favipiravir** | **Ciclesonide** | **Heparin** | **Tocilizumab** |
| --- | --- | --- | --- | --- | --- | --- |
| **No comorbidity** | 1873 | 105 (6) | 606 (32) | 562 (30) | 48 (3) | 22 (1) |
| **Obesity** | 178 | 34 (19) | 109 (61) | 72 (40) | 17 (10) | 5 (3) |
| **Cardiovascular disease** | 183 | 30 (16) | 86 (47) | 46 (25) | 19 (10) | 6 (3) |
| **Chronic respiratory disease** | 140 | 39 (28) | 94 (67) | 52 (37) | 10 (7) | 5 (4) |
| **Diabetes** | 562 | 98 (17) | 325 (58) | 197 (35) | 61 (11) | 12 (2) |
| **Hypertension** | 623 | 120 (19) | 338 (54) | 200 (32) | 70 (11) | 20 (3) |
| **Cerebrovascular disease** | 187 | 34 (18) | 93 (50) | 42 (23) | 15 (8) | 7 (4) |
| **Liver disease** | 88 | 15 (17) | 57 (65) | 32 (36) | 5 (6) | 3 (3) |
| **Severe renal disease or dialysis** | 47 | 9 (19) | 21 (45) | 12 (26) | 6 (13) | 3 (6) |
| **Solid tumor** | 145 | 20 (14) | 68 (47) | 34 (23) | 6 (4) | 1 (1) |

**Supplementary Figure 1.**
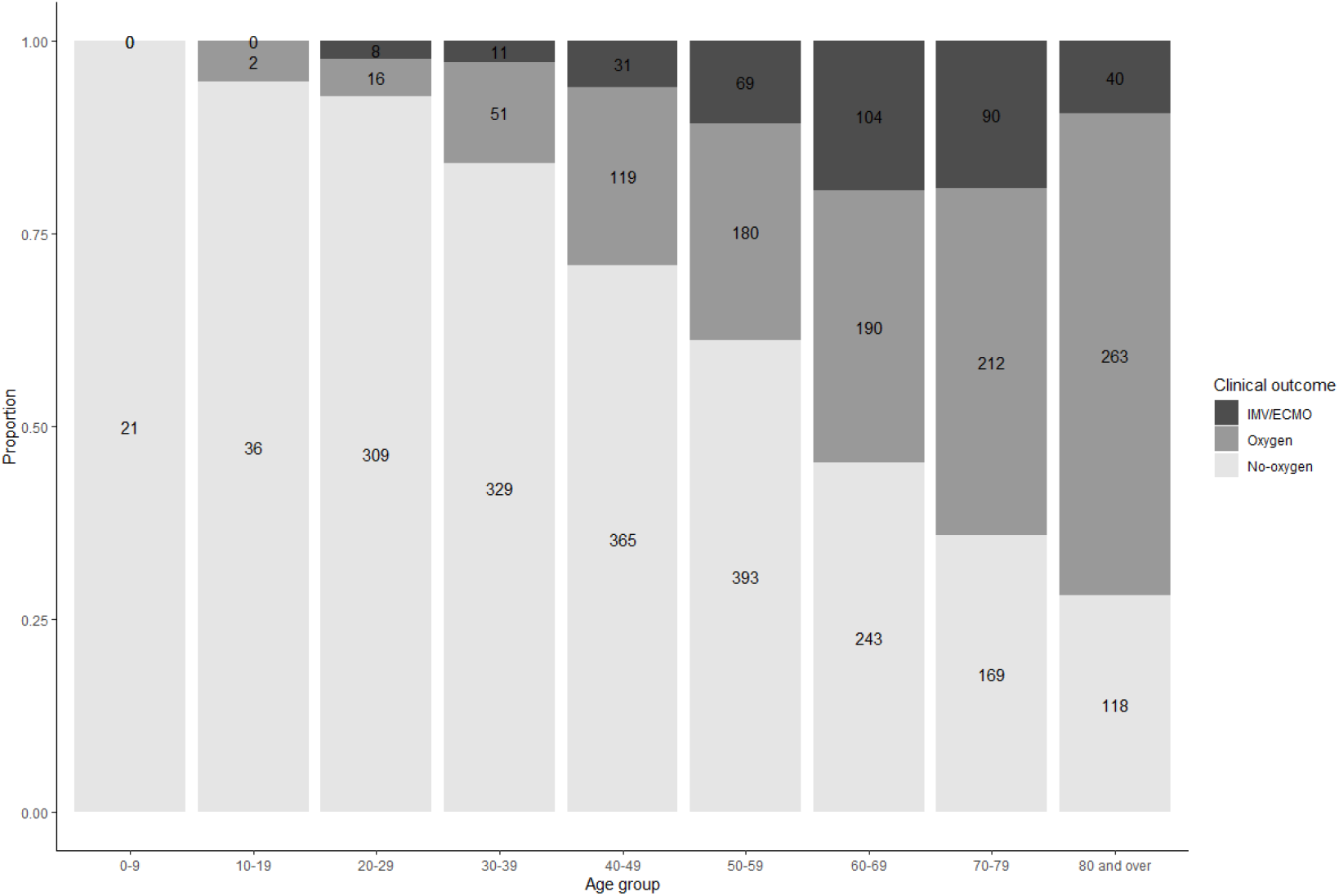
Distribution of worst severity by age group.

**Supplementary Figure 2.**
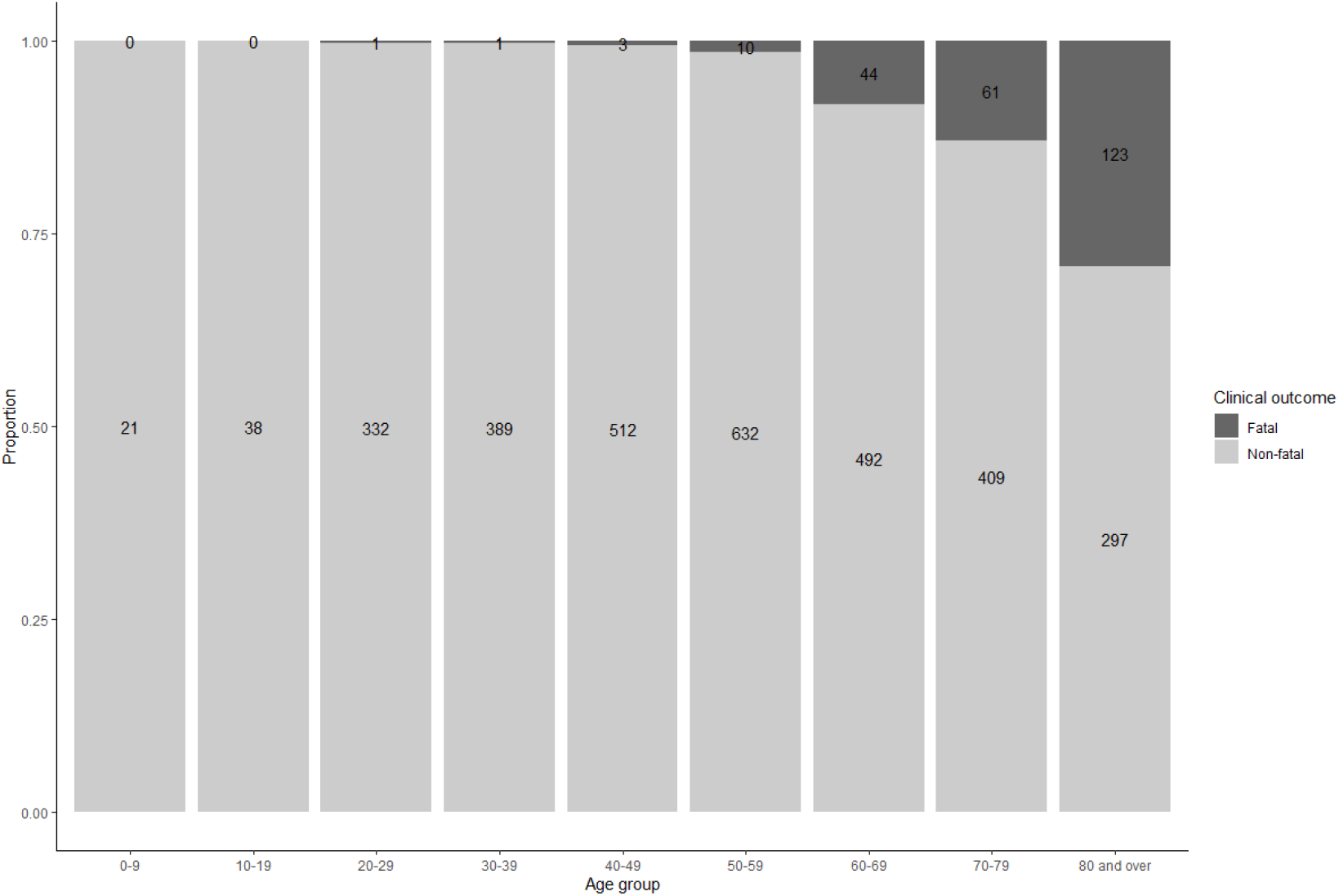
Distribution of fatality by age group.

**Supplementary Figure 3.**
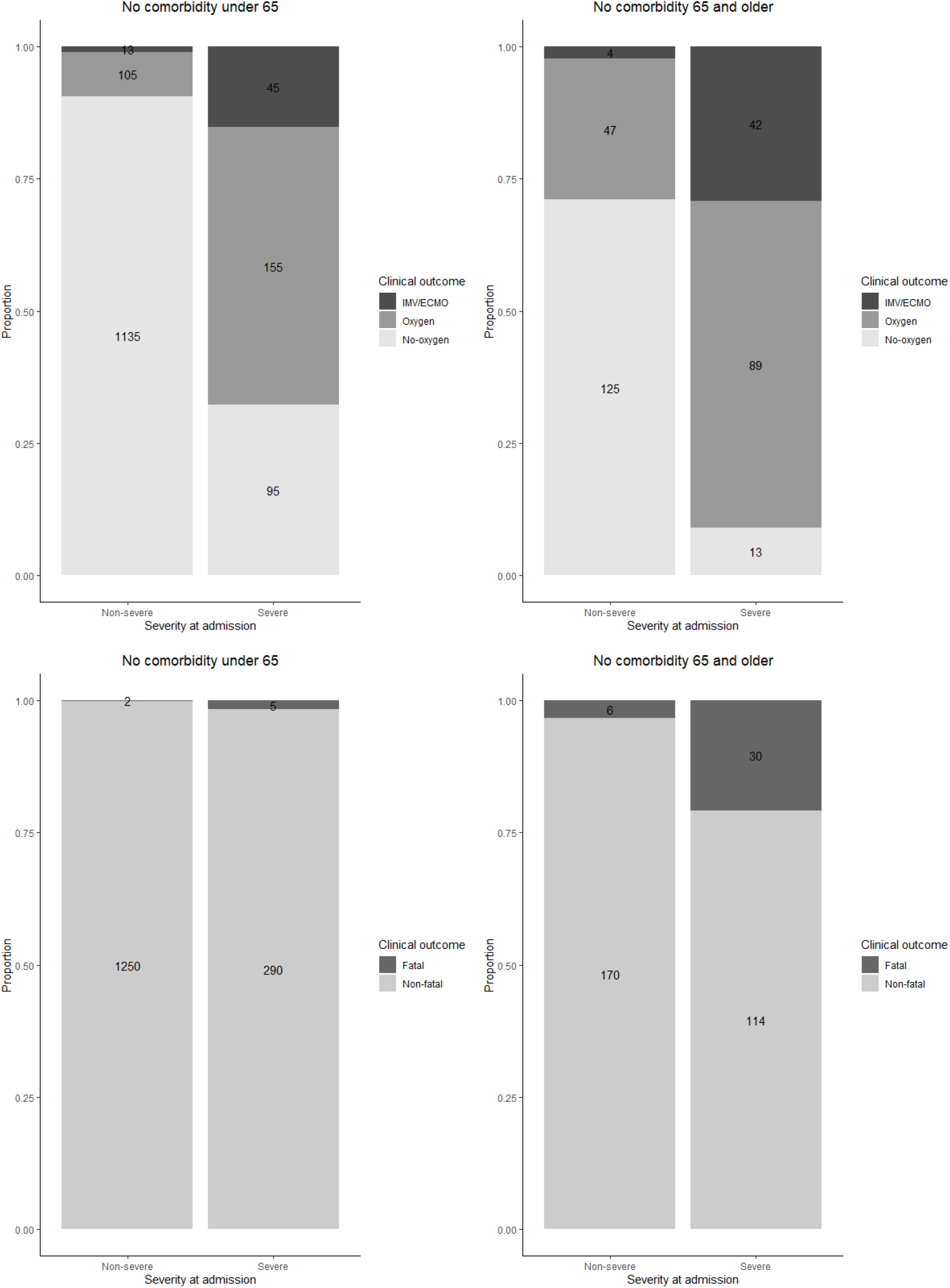
(a) Worst severity of cases aged <65 and ≥65 with no comorbidities (b) Fatality in cases aged <65 and ≥65 with no comorbidities.

